# Mortality and associated factors among hospitalized COVID 19 patients in Lira regional referral hospital, a cross sectional study

**DOI:** 10.1101/2024.10.05.24314935

**Authors:** Ongoli Augustine, Opio Bosco, Marc Sam Opollo, Ann Ruth Akello, Kiweewa Francis, Omech Bernard

**Affiliations:** Department of Public Health Lira University; Uganda Christian Institute For Nursing and Midwifery; Department of Internal Medicine Lira Regional Referral Hospital; Strengthening Institutional Capacity for Research Administration

**Keywords:** COVID 19, Mortality, COVID 19 Treatment Unit (CTU), Lira Regional Referral Hospital (LRRH), Uganda, All three waves, Between May 2020 and March 2022

## Abstract

**Background:** Corona Virus Disease 2019 (COVID-19) caused public emergency with serious morbidity and mortality worldwide between 2020 and 2022, The direct impact of the disease and associated mortality may have been relatively limited in Sub-Saharan Africa (SSA), compared to the impact in other regions. The factors that are considered high risk for acquisition of COVID-19 and associated high mortality rate amongst in-patients are varied in different settings, there are limited data from regional referral designated COVID-19 Treatment Units (CTU) in Uganda. This research assessed the mortality rate, the sub-populations at high risk and characteristics of COVID-19 patients hospitalized in Lira Regional Referral Hospital (LRRH).

**Objective:** To describe COVID-19 characteristic, mortality and associated risk factors among patients admitted at the Lira Regional referral hospital COVID 19 Treatment Unit in northern Uganda.

**Design:** Cross Sectional Study with use of Secondary Data

**Setting:** This study was conducted at Lira RRH between January 2023 and December 2023. The data used were for patients admitted from May 2020 to March 2022.

**Participant:** In this study all the patients with confirmed COVID-19 were selected by simple census sampling technique, 490 participants were included in the study and these were a) moderately to critically ill patients, b) mild or asymptomatic patients with comorbidity, c) those with positive COVID-19 test and d) those who were admitted in the hospital.

**Results:** In the final analysis, 490 participants were considered. Out of this, 251 (52%) were females. Majority 203(41%) were older than 60 years of age. Most of the patients presented with cough 369(89.56%), difficulty in breathing (DIB) 293(78.76%), chest pain 237(69.3%), general body weakness (GBW) 199(63.38%) and fever 179(61.3%). Common pre-existing comorbidities were hypertension 139(29.96%), diabetes mellitus 89(19.47%) and HIV 44(10%). Of all the patients admitted, 187(40%) had severe disease and 34(7%) were critically ill.

Overall from May 2020 to March 2022, 142(29%) died. Oxygen saturation (SPO2) 92-100% had 89% decreased mortality (aOR-0.11, 95% CI 0.03-0.44, p-value-0.002). Body temperature 35.5-37.5 degrees Celsius had 78% decreased mortality (aOR-0.22, 95% CI 0.05-0.99, p-value-0.049). Those without Chronic Liver Disease (CLD) had 99% decreased mortality (aOR-0.01, 95% CI 0.001-0.46, p-value-0.017). Age 31-45yrs had 86% decreased mortality (aOR-0.14, 95% CI 0.03-0.74, p-value-0.021)

**Conclusion:** The in-hospital mortality rate in our cohort of COVID 19 patients admitted at LRRH was high. Not having chronic liver disease, normal oxygen saturation, normal body temperature and younger age were associated with decreased likelihood of death.

## Background

Corona Virus Disease 2019 (COVID 19) is a respiratory disease caused by the severe acute respiratory syndrome corona virus type 2 (SARS Cov 2), which caused a global pandemic with substantial morbidity and mortality between 2020 and 2022 [1]. The overall pooled Case Fatality Rate (CFR) for COVID 19 globally was 10.0% in 2020 when mortality was highest but with different CFR in different settings e.g. 1% in the general population, 15% in hospitalized patients and 29% in ICU patients [2]. There was a higher CFR for those with co-morbidities such cardiovascular illnesses-13.2%, diabetes mellitus (DM)-9.2%, high blood pressure (HTN)-8.4%, respiratory disorders-8.0%, and cancer-7.6% [3]. The CFR has since then been dropping globally to less than 3% by August 2022 [4]. Sub-Saharan Africa (SSA) had a CFR of about 3.0% [5]. There are few recommended treatments for COVID 19 like Nirmatrelvir-Ritonavir and Remdesivir for mild to moderate cases, Baricitinib and Tocilizumab for severe cases in hospitalized settings [6] [7] [8] [9]. However the world has embarked vaccination and other preventative measures such as social isolation, masking, frequent hand washing, and social distancing as the cornerstone of management [10].

It has been observed that 75% of the people hospitalized with COVID 19 have at least one associated risk factor [11]. The risk of death is more marked in people with certain risk factors like the elderly, pregnancy, male gender and people with underlying medical conditions (like cancer, hypertension, diabetes, sickle cell anemia, kidney disease, liver disease, chronic lung diseases, etc) [12] [13]. COVID 19 outcomes like death and post acute sequel of COVID (PASC) are affected by the above mentioned risk factors.

Some studies have found out that these factors do not affect COVID19 outcomes e.g. HIV does not affect COVID 19 outcome[14]. Another study also reported that women were more affected by COVID 19 than men [15]. This study intended to understand the scenario of COVID 19 and the associated factors in Lira Regional Referral in northern Uganda.

### Main Objective

To determine the mortality rate, describe the clinical characteristics, factors associated with mortality of patients who were admitted with COVID 19 in the CTU at Lira regional referral hospital

### Specific objectives

1) To determine the mortality rate of the patients hospitalized in the CTU at LRRH between MAY 2020 and April 2022 2) To describe the clinical characteristic of COVID 19 patients admitted in CTU at LRRH between May 2020 to April 2022 3) To establish the factors associated with mortality among COVID 19 patients hospitalized in CTU at LRRH between May 2020 and April 2022.

## Methods

### Study Design

We conducted a cross sectional study using secondary data of patients admitted at Lira Regional Referral Hospital (LRRH) between May 2020 and March 2022.

### Study settings

The study was conducted at LRRH COVID 19 Treatment Unit (CTU) between January 2023 and December 2023. LRRH is located in Northern Uganda, Lira city, Lira city east division, near Lira central police station and it serves 9 districts of the region. LRRH-CTU is a 30 bed capacity and they admitted a total of 795 patients between May 2020 and April 2022

#### Target population

Patients suffering from COVID 19.

#### Study population

COVID 19 patients who are admitted at Lira Regional Referral Hospital.

#### Inclusion Criteria

a) patients with positive COVID 19 test either by Polymarase Chain Reaction Reverse Transcriptase (PCR-RT) Test or by Covid Rapid Diagnostic Test (CRDT), b) Patients 18years and above, c) Patients admitted in the CTU, d) Symptomatic patients, e) Patients with comorbidity.

#### Exclusion criteria

a) Charts missing more than 75% of the predefined information in the data abstraction tool, b) charts with unclear bad handwriting that cannot allow easy reading and comprehension, c) Asymptomatic patients without comorbidity.

### Data Collection

All the files were retrieved from the archive at records by the PI and each file assessed for eligibility. Data was collected on the patient’s bio-data (Age, sex, religion, occupation, tribe), clinical signs and symptoms (blood pressure, pulse rate, oxygen saturation, temperature, cough, fever, myalgia, malaise, dyspnea and others), underlying medical conditions (Cancer, Hypertension, HIV, DM, Malaria, Pregnancy and others), grades of disease severity (asymptomatic, mild, moderate, severe and critical), treatment given and outcome status (alive and died) and recorded. Our outcome of interest was death and recovered. All measures were taken to ensure strict confidentiality of the data collected.

### Data Management and Analysis

The anonymized data abstracted from the patient’s charts were entered in STATA for analysis. All data were coded and then entered in a database that have logic checks to ensure that mistakes were identified as data is entered and progression was only enabled after these were corrected.

Categorical data, the status of the patients at discharge (dead or recovered) were presented as Frequencies and Percentages. Continuous data were described using Mean and standard deviation (S.D). The independent associations between the dependent variables (recovered and died) and the independent variable (Age, Sex, HIV, Tuberculosis, Hypertension, SCD, Pregnancy, Malaria, cancer, Asthma, COPD, Liver cirrhosis, Hepatitis B, Chronic renal disease, Cerebral Vascular disease, Diabetes mellitus type 1 and 2, Obesity, Smoking, use of corticosteroids), was tested using Chi Square and the Chi Square value reported.

A bivariate logistic regression was done determine the risk factors (predictors) for mortality among COVID 19 patients hospitalized in CTU and the results were reported as crude odds ratio (cOR) and a multivariate logistic regression was also done and results reported as adjusted odds ratio (aOR). Test for significance was done at 95% confidence interval.

### Ethical Considerations

This research was approved by Gulu research ethics committee (GUREC-2022-357), the study was conducted according to the principles of the Helsinki declaration.

#### Reporting

We used the STROBE cross sectional reporting guidelines for this study [16]

## Results

### Recruitment and selection

Out of the 795 patients who were admitted in LRRH-CTU by March 2022, we extracted data from 490 patient charts. 150 patients below 18years and were excluded. 25 files with less than 75% of the stipulated information the data abstraction form excluded. 130 patients who were asymptomatic and did not have any underlying comorbidity were also excluded. See Table 1.

**Table 1.**
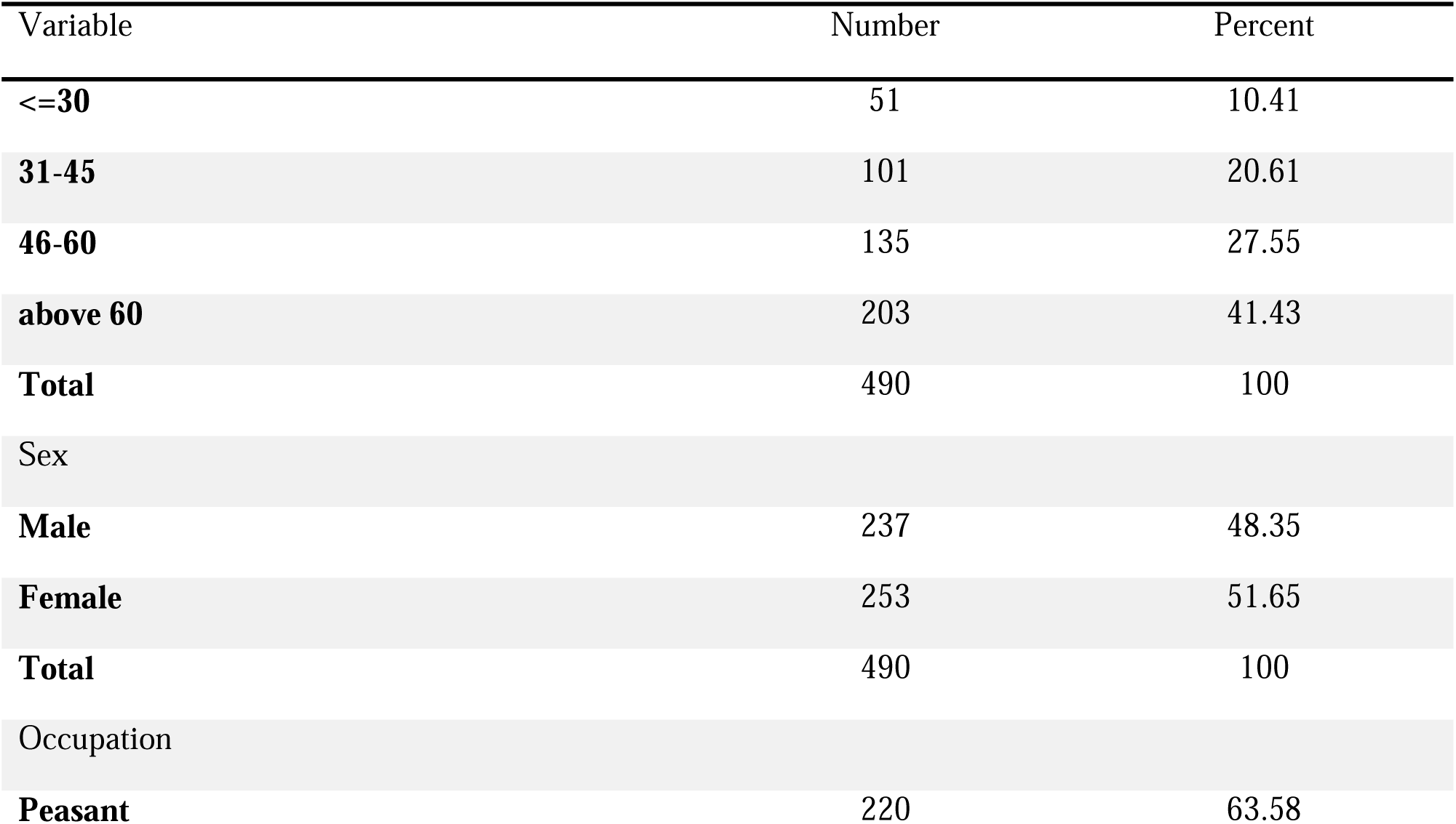
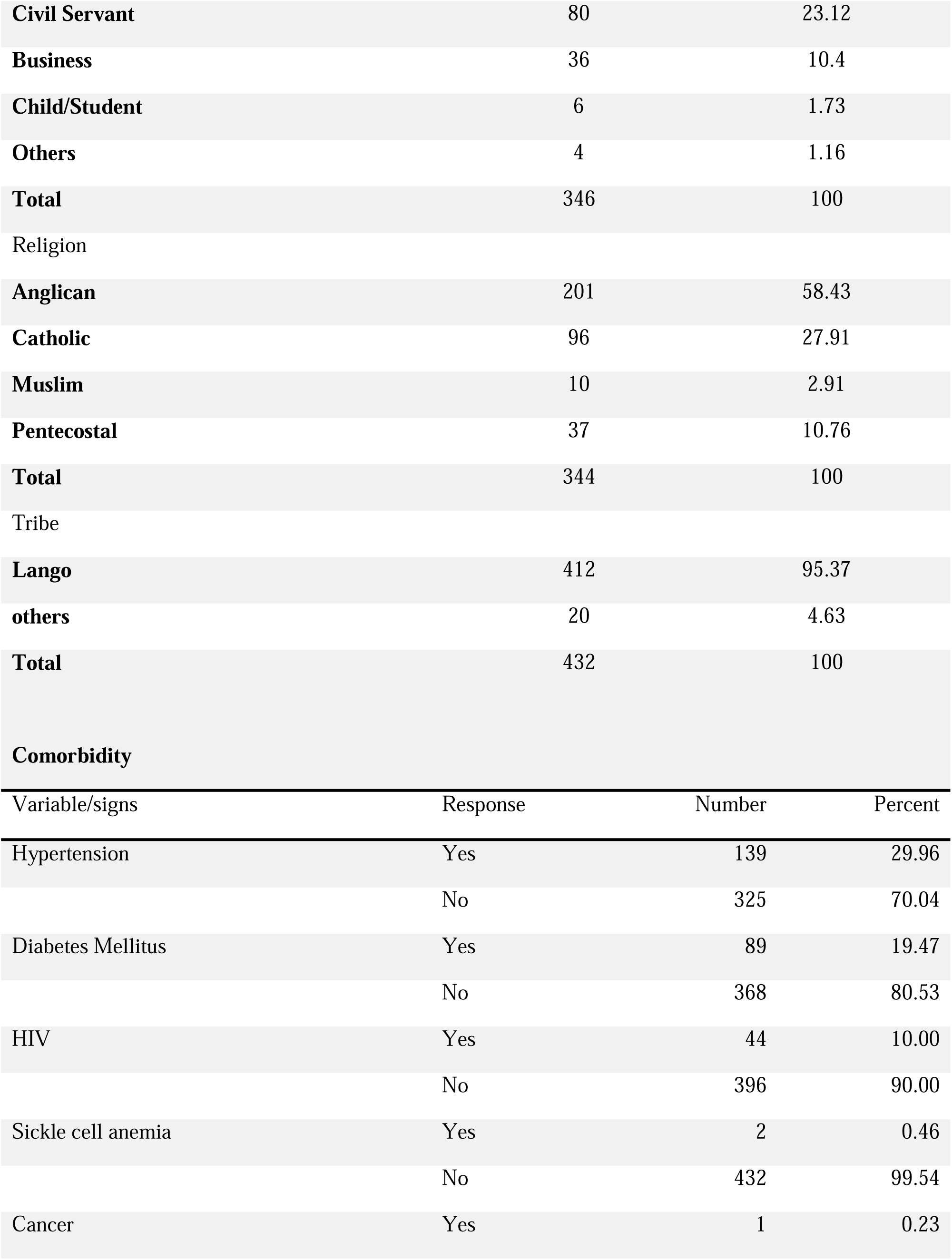
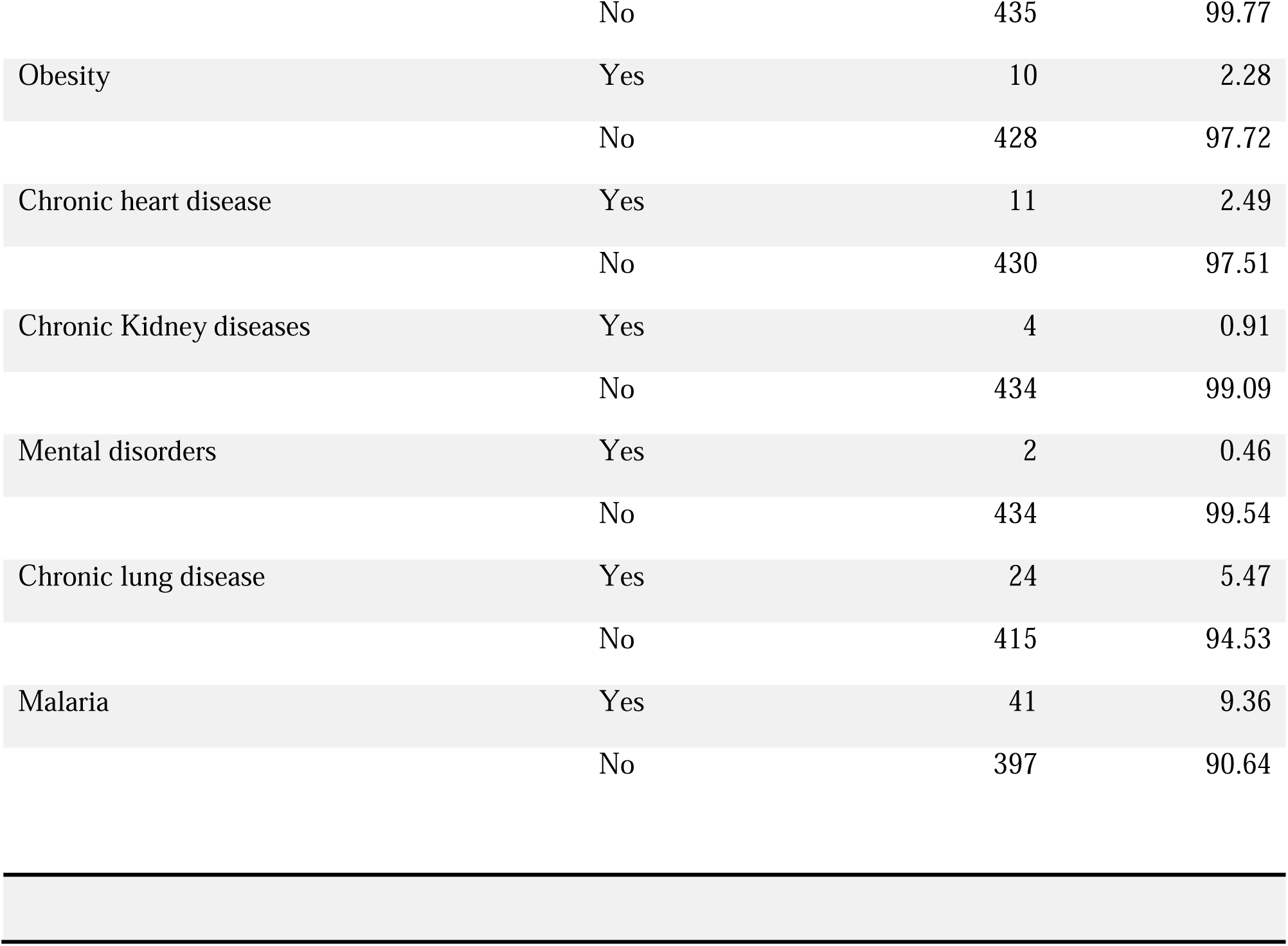
Social demograhic characteristic and comorbidity.

| Variable | Number | Percent |
| --- | --- | --- |
| **<=30** | 51 | 10.41 |
| **31-45** | 101 | 20.61 |
| **46-60** | 135 | 27.55 |
| **above 60** | 203 | 41.43 |
| **Total** | 490 | 100 |
| Sex |  |  |
| **Male** | 237 | 48.35 |
| **Female** | 253 | 51.65 |
| **Total** | 490 | 100 |
| Occupation |  |  |
| **Peasant** | 220 | 63.58 |
| **Civil Servant** | 80 | 23.12 |
| **Business** | 36 | 10.4 |
| **Child/Student** | 6 | 1.73 |
| **Others** | 4 | 1.16 |
| **Total** | 346 | 100 |
| Religion |  |  |
| **Anglican** | 201 | 58.43 |
| **Catholic** | 96 | 27.91 |
| **Muslim** | 10 | 2.91 |
| **Pentecostal** | 37 | 10.76 |
| **Total** | 344 | 100 |
| Tribe |  |  |
| **Lango** | 412 | 95.37 |
| **others** | 20 | 4.63 |
| **Total** | 432 | 100 |

Comorbidity
| Variable/signs | Response | Number | Percent |
| --- | --- | --- | --- |
| Hypertension | Yes | 139 | 29.96 |
|  | No | 325 | 70.04 |
| Diabetes Mellitus | Yes | 89 | 19.47 |
|  | No | 368 | 80.53 |
| HIV | Yes | 44 | 10.00 |
|  | No | 396 | 90.00 |
| Sickle cell anemia | Yes | 2 | 0.46 |
|  | No | 432 | 99.54 |
| Cancer | Yes | 1 | 0.23 |
|  | No | 435 | 99.77 |
| Obesity | Yes | 10 | 2.28 |
|  | No | 428 | 97.72 |
| Chronic heart disease | Yes | 11 | 2.49 |
|  | No | 430 | 97.51 |
| Chronic Kidney diseases | Yes | 4 | 0.91 |
|  | No | 434 | 99.09 |
| Mental disorders | Yes | 2 | 0.46 |
|  | No | 434 | 99.54 |
| Chronic lung disease | Yes | 24 | 5.47 |
|  | No | 415 | 94.53 |
| Malaria | Yes | 41 | 9.36 |
|  | No | 397 | 90.64 |

### Social demographic characteristics and comorbidity

Table 1 summarizes the social demographic characteristics and comorbidity of patients admitted in LRRH CTU. Overall, the majority of patients admitted with COVID 19 at LRRH (69%) were 46 years or older, with those aged 60 year or older making up 41.33%. Most of the patients admitted to the CTU were females 253(51.65%). Most commonly patients presented with hypertension 139(29.96%), diabetes mellitus 89(19.47%), HIV 44(10%). See Table 1

### Clinical presentation

Most patients presented to the CTU with cough 369(89.56%), difficulty in breathing (DIB) 293(78.76%), chest pain 237(69.3%), general body weakness (GBW) 199(63.38%) and fever 179(61.3%). At least 179(47.73%) presented with low oxygen saturation SPO2**<**92%, this was the lowest oxygen saturation recorded throughout the disease course during hospitalization of all patients. See Table 2

**Table 2.**
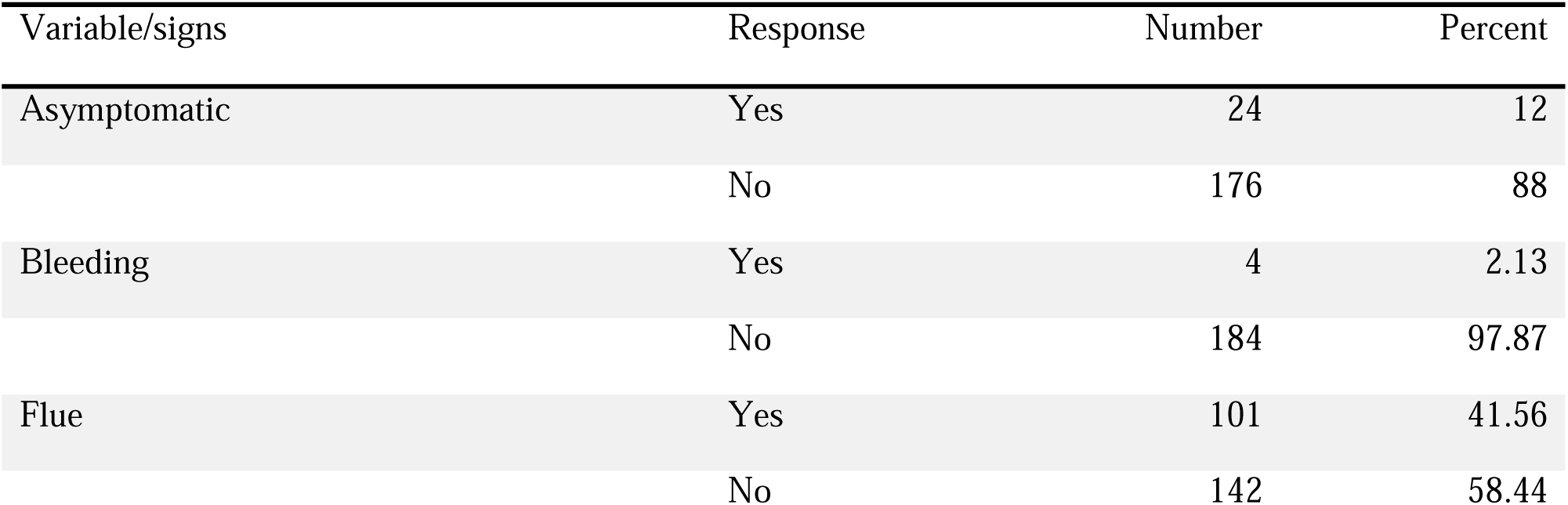
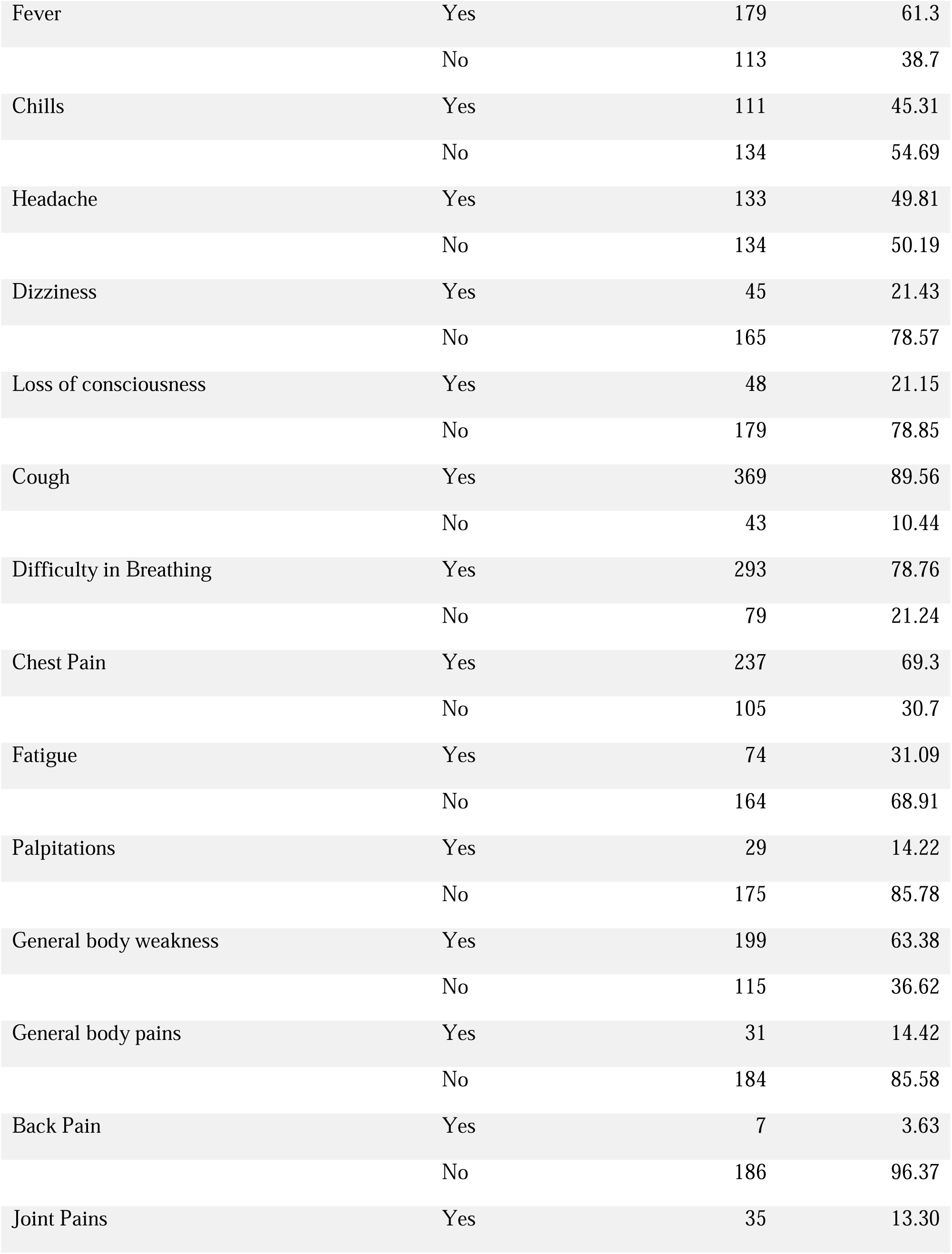
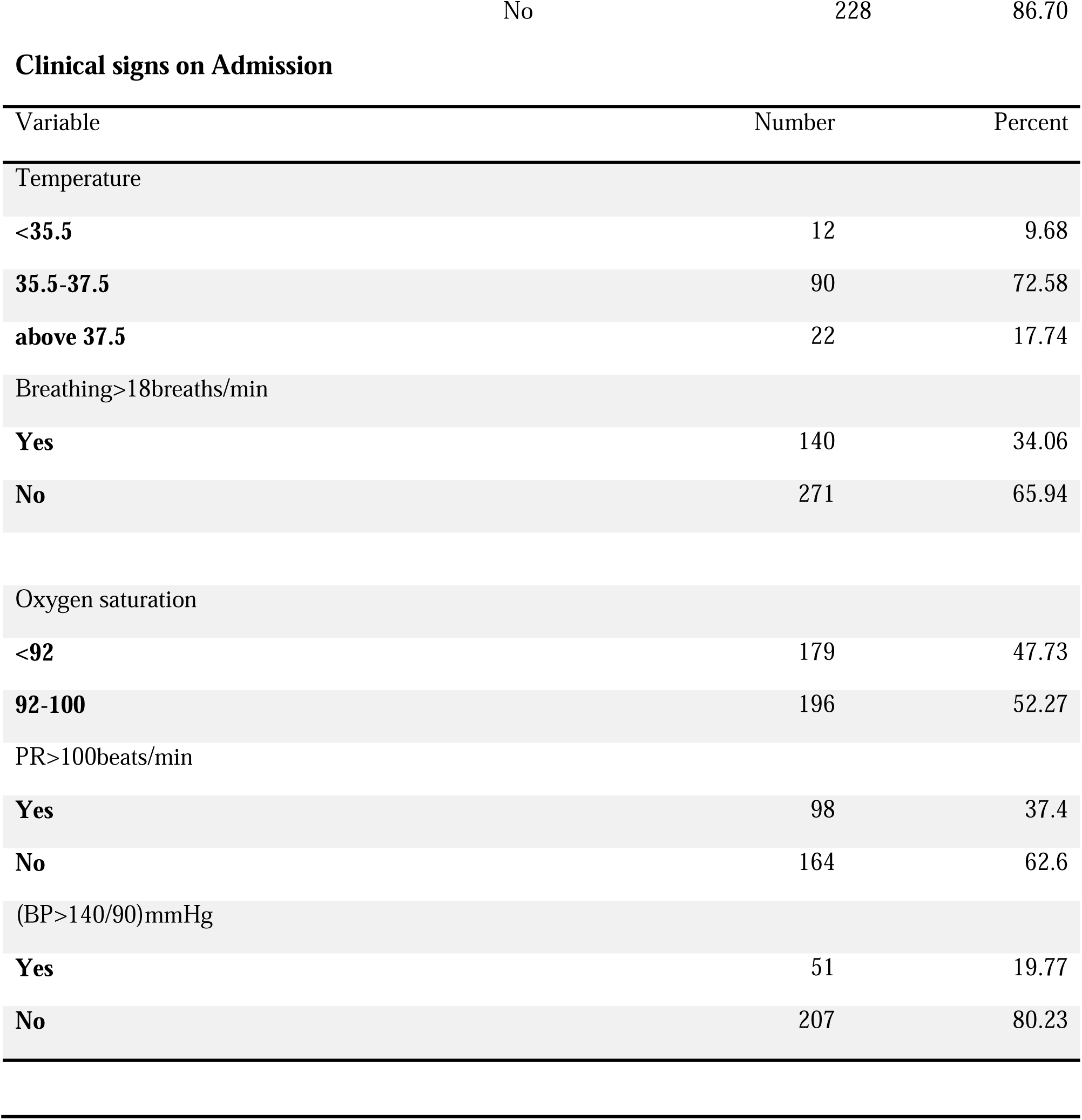
Common symptoms throughout the disease course and clinical signs on admission. However the lowest oxygen saturation throughout the disease course was the one taken.

### Treatment given

Most patients 356(73.4%) received antibiotic therapy. Many patients 221(48.39%) presented with severe and critical disease. See Table 3.

**Table 3.**
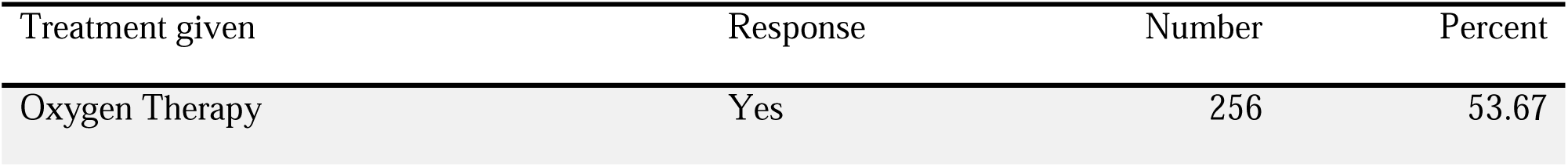
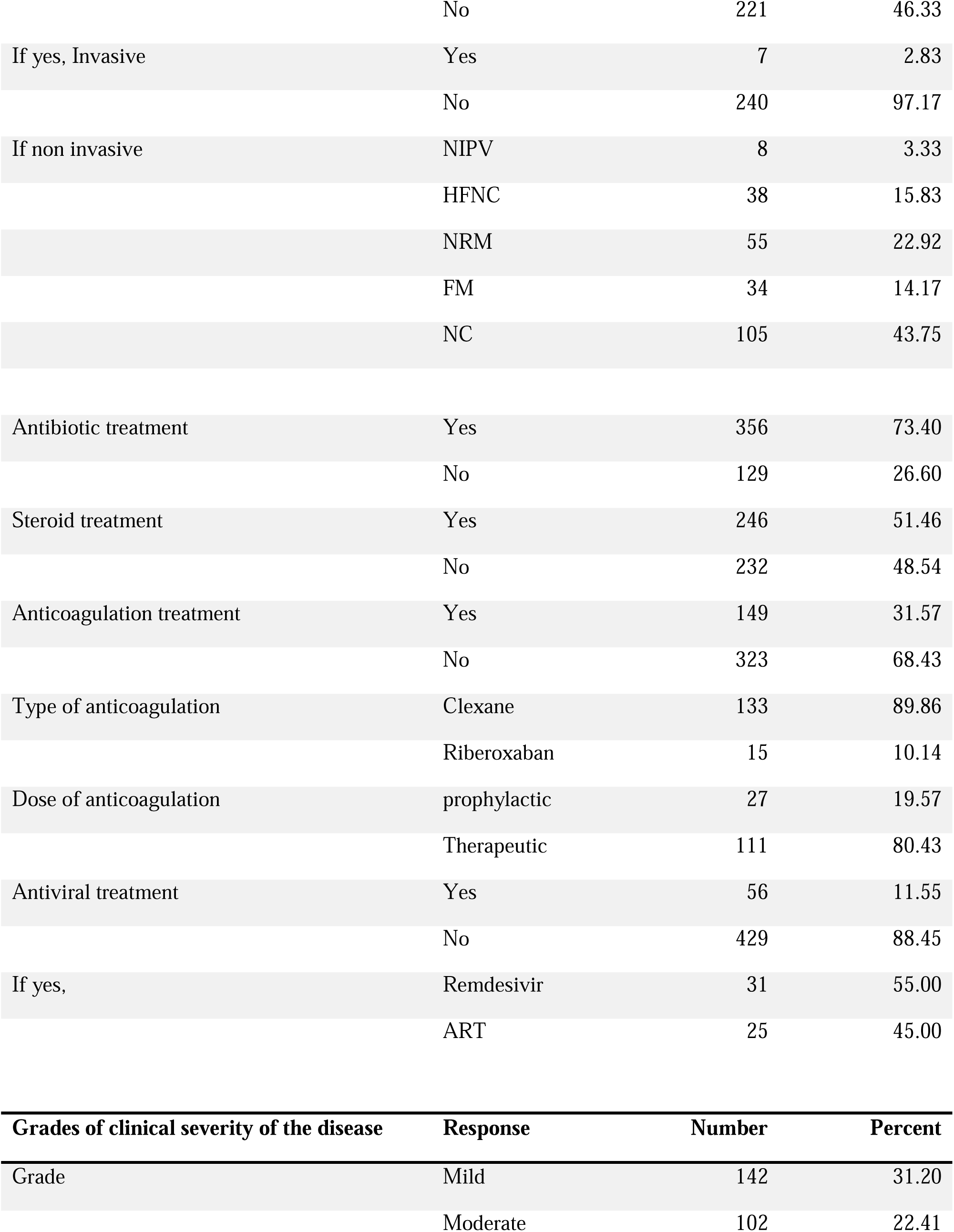
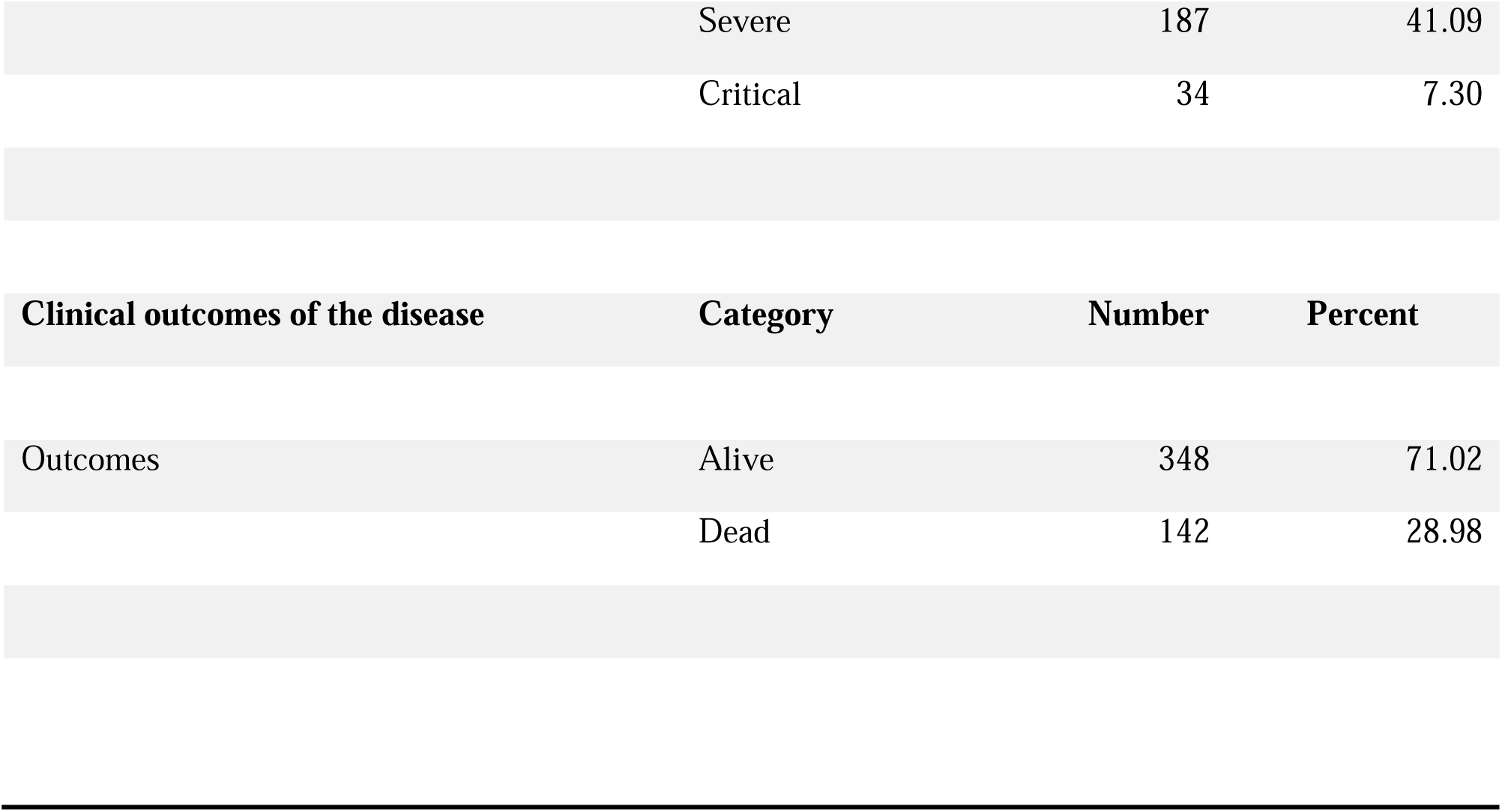
Treatment given, grades of disease severity and clinical outcomes of the disease.

### Bivariate analysis of risk factors

A bivariate logistic regression was conducted, statistical significance was suggested if the p-value < 0.05 and crude odds ratio (cOR) > 1 at 95% confidence interval.

Age above 60 years was 6 times likely to die (COR-6.47, CI 2.64-15.8, p-value < 0.001). Females had a decreased mortality rate (COR-0.78, CI 0.53-0.61, p-value < 0.001).

People with normal oxygen saturation (SPO2 92% to 100%) had decreased mortality rate (COR-0.21, CI 0.13-0.33, p-value < 0.001).

Normal respiratory rate (12-18) breaths per minute was associated with decreased mortality rate (COR-0.19, CI 0.12-0.32, p-value < 0.001).

People who did not receive oxygen had a lower mortality rate (COR-0.12, CI 0.07-0.21, p-value < 0.001)

People who were not treated with steroids had a decreased mortality rate (COR-0.23, CI 0.15-0.37, p-value < 0.001).

Patients without DM had decreased mortality rate (COR-0.41, CI 0.25-0.67, p-value < 0.01) Patients without hypertension were less likely to die (COR-0.46, CI 0.29-0.71, p-value = 0.001). See Table 4

**Table 4.** Bivariate analysis and Multivariate Analysis.

| Death |  |  |  |  |  |
| --- | --- | --- | --- | --- | --- |
| Variables |  | Yes(1) | No (0) | COR (95%CI) | p value |
| Age | <30 | 6(11.76) | 45(88.24) | 1.00 |  |
|  | 31-45 | 12(12.00) | 88(88.00) | 1.02(0.36-2.90) | 0.966 |
|  | 46-60 | 28(20.9) | 106(79.10) | 1.98(0.77-5.11) | 0.158 |
|  | Above 60 | 94(46.31) | 109(53.69) | 6.47(2.64-15.8) | <0.001* |
| Sex | Male | 74(31.5) | 161(68.5) | 1.00 |  |
|  | Female | 66(26.5) | 183(73.5) | 0.78(0.53-0.61) | <0.001* |
| Oxygen saturation | <92 | 100(55.87) | 79(44.13) | 1.00 |  |
|  | 92-100 | 41(21.13) | 153(78.87) | 0.21(0.13-0.33) | <0.001* |
| Temperature | <35.5 | 7(58.33) | 5(41.67) | 1.00 |  |
|  | 35.5-37.5 | 30(33.71) | 59(66.29) | 0.36(0.11-1.24) | 0.106 |
|  | > 37.5 | 9(40.91) | 13(59.09) | 0.49(0.12-2.06) | 0.334 |
| Breathing >18breath/min | Yes | 63(45.0) | 77(55.0) | 1.00 |  |
|  | No | 37(13.75) | 232(86.25) | 0.19(0.12-0.32) | <0.001* |
| Tachycardia | Yes | 45(45.92) | 53(54.08) | 1.00 |  |
|  | No | 50(30.86) | 112(69.14) | 0.53(0.31-0.88) | 0.015* |
| Chronic heart disease | Yes | 6(54.55) | 5(45.45) | 1.00 |  |
|  | No | 94(21.96) | 334(78.04) | 0.23(0.07-0.78) | 0.019* |
| Chronic liver disease | Yes | 5(62.5) | 3(37.5) | 1.00 |  |
|  | No | 93(21.78) | 334(78.22) | 0.17(0.04-0.71) | 0.016* |
| Chronic neurological disorder | Yes | 7(58.33) | 5(41.67) | 1.00 |  |
|  | No | 94(22.17) | 330(77.83) | 0.20(0.06-0.66) | 0.008* |
| Current acute febrile illness | Yes | 17(42.50) | 23(57.50) | 1.00 |  |
|  | No | 83(20.96) | 313(79.04) | 0.36(0.18-0.70) | 0.003* |
| Smoking status | Yes | 10(38.46) | 16(61.54) | 1.00 |  |
|  | No | 111(25.28) | 328(74.72) | 0.54(0.24-1.23) | 0.142 |
| Alcohol use | Yes | 10(41.67) | 14(58.33) | 1.00 |  |
|  | No | 109(24.77) | 331(75.23) | 0.46(0.20-1.10) | 0.071 |
| Received O2 therapy | Yes | 117(45.70) | 139(54.30) | 1.00 |  |
|  | No | 21(9.50) | 200(90.50) | 0.12(0.07-0.21) | <0.001* |
| Steroid treatment received | Yes | 105(42.68) | 141(57.32) | 1.00 |  |
|  | No | 35(15.09) | 197(84.91) | 0.23(0.15-0.37) | <0.001* |
| Hypertension | Yes | 51(36.96) | 87(63.04) | 1.00 |  |
|  | No | 69(21.30) | 255(78.70) | 0.46(0.29-0.71) | 0.001* |
| Diabetes | Yes | 35(40.23) | 52(59.77) | 1.00 |  |
|  | No | 79(21.47) | 289(78.53) | 0.41(0.25-0.67) | <0.001* |
| Received ARV | Yes | 10(17.86) | 46(82.14) | 1.00 |  |
|  | No | 130(30.30) | 299(69.70) | 2.00(0.97-4.08) | 0.057 |

\* significant

Multivariate Analysis
|  |  | Death |  | AOR (95%CI) | p value |
| --- | --- | --- | --- | --- | --- |
| Variables |  | Yes(1) | No (0) |  |  |
| Age (years) | <30 | 6(11.76) | 45(88.24) | 1.00 |  |
|  | 31-45 | 12(12.00) | 88(88.00) | 0.14(0.03-0.74) | 0.021* |
|  | 46-60 | 28(20.9) | 106(79.10) | 0.75(0.19-2.94) | 0.685 |
|  | Above 60 | 94(46.31) | 109(53.69) | - | - |
| Sex | Male | 74(31.5) | 161(68.5) | 1.00 |  |
|  | Female | 66(26.5) | 183(73.5) | 0.34(0.08-1.45) | 0.145 |
| Oxygen saturation | <92 | 100(55.87) | 79(44.13) | 1.00 |  |
|  | 92-100 | 41(21.13) | 153(78.87) | 0.11(0.03-0.44) | 0.002* |
| Temperature (<sup>0</sup>C) | <35.5 | 7(58.33) | 5(41.67) | 1.00 |  |
|  | 35.5-37.5 | 30(33.71) | 59(66.29) | 0.22(0.05-0.99) | 0.049* |
|  | > 37.5 | 9(40.91) | 13(59.09) | 0.36(0.04-3.74) | 0.396 |
| Breathing >18breath/min | Yes | 63(45.0) | 77(55.0) | 1.00 |  |
|  | No | 37(13.75) | 232(86.25) | 3.03(0.68-13.35) | 0.148 |
| Chronic liver disease | Yes | 5(62.5) | 3(37.5) | 1.00 |  |
|  | No | 93(21.78) | 334(78.22) | 0.01(0.001-0.46) | 0.017* |
| Received O<sub>2</sub> therapy | Yes | 117(45.70) | 139(54.30) | 1.00 |  |
|  | No | 21(9.50) | 200(90.50) | 0.26(0.06-1.08) | 0.064 |
| Steroid treatment received | Yes | 105(42.68) | 141(57.32) | 1.00 |  |
|  | No | 35(15.09) | 197(84.91) | 0.76(0.20-2.86) | 0.680 |

### Multivariate analysis of data shows

Multivariate logistic regression was conducted, statistical significance was suggested if the p-value < 0.05 and crude odds ratio (cOR) > 1 at 95% confidence interval.

Normal oxygen saturation was associated with decreased mortality rate compared to low oxygen saturation (aOR-0.11, CI 0.03-0.44, p-value = 0.002).

Normal body temperature was associated to low mortality rate compared to abnormal body temperatures, (aOR-0.22, CI 0.05-0.99, p-value = 0.049).

Patients without chronic liver disease had a decreased mortality rate compared to patients with chronic liver disease (aOR-0.01, CI 0.001-0.46, p-value = 0.017). See table 4.

## Discussion

### Clinical outcomes

Of all the patients admitted in the CTU, 142 (29%) died during admission. This finding is higher than that reported Gulu regional referral hospital (GRRH) 4.8 % between march 2020 and October 2021 [17], the low mortality rate in GRRH could have been because their study included patients aged 12 years and above and those with asymptomatic disease. Similarly, a report from a private hospital in kampala found 11% mortality during the June 2020 and September 2021 [18], However, our findings is lower than that reported at Mulago National Referral Hospital with a mortality rate of 37% during the second wave (between May 1 and July 11, 202) [15], This could be because of the selective nature of their research that was done only during the second wave and done only on patients in High Dependency Unit (HDU) that are very sick. The wide variations of COVID 19 mortality in Uganda designated CTUs could be largely explained by the difference in population characteristics and the period under considerations. There were three waves of COVID-19 pandemic in Uganda including delta variant which was the most fatal and dominated the second wave and Omicron which was the most infectious and dominated the third wave [19] [20]. Our study covered the entire period of the epidemic in Uganda and yet the mortality could have peaked during one major wave, it also excluded patients who were hospitalized with asymptomatic diseases merely for isolation or home based care as well as children below 18 years of age. Of all the patients who were admitted to the CTU, 31% had mild diseases, 22% had moderate disease, 40% had severe disease and 7% had critical disease. The high mortality rate (29%) could be attributed to the fact that many patients admitted in the CTU had severe cases of COVID 19 who were more likely to die. Notwithstanding, the availability and utilization of lifesaving interventions at the CTU at the facility at the time may have impacted on the outcomes of the patients admitted at CTU at LRRH.

### Factors associated with mortality

The admission rate were 52% for female and 48% for males, this could be because women are naturally fearful and generally have better health seeking behavior compared to males, however sex was not found to be associated with mortality as opposed to other studies [21] [22].

Majorities of those admitted to the CTU (41%) were individuals over 60 years. Age 45 years and below was associated with 86% decreased mortality (aOR-0.14, 95% CI 0.03-0.74, p-value-0.021). This could be because this category of people is immunocompetent and most of them do not have associated comorbid diseases, our finding is consistent with other studies [23][24]

Most patients (90%) presented to the CTU with cough, DIB (79%), chest pain (69%), GBW (63%) and fever (61%). These are the most common and more specific signs of COVID 19. The least presenting symptoms were back pain (4%) and bleeding (2%) are rare and least specific symptoms of COVID 19. Ageusia and Anosmia are some rare symptoms of COVID 19 with symptom sensitivities of less than 50% but almost everyone who presented with them without any other system disease that explains them tested positive for COVID 19, symptom specificities of over 90% [25]. There were 12% asymptomatic patients admitted in the CTU, this is because some patients with comorbidity and positive COVID 19 test were admitted for close monitoring but progressed to seroconvert without developing signs and symptoms were also included in this study. Most patients (72%) presented with temperature within the normal range 35.5 to 37.5, few patients (18%) were hypothermic <35.5 and others (18%) had hyperthermia >37.5. about the number of patients with hyperthermia were far less compared to the number of patients with reported history of fever probably because this study took record of vital signs (Temp) at admission only yet it recorded symptoms (Fever) at any time of the disease course. Often times patients report fever in COVID 19 without associated high body temperature, this is consistent with a study done in china where 1099 patients were studied and only 44% had temp > 37.5 degrees Celsius on admission but ultimately was noted in 89% during the period of hospitalization [26]. There is paucity of data on the pattern of fever in COVID 19 and this was beyond the scoop of this study. Patients who presented with temperature within normal range (35.5 degrees Celsius to 37.5 degrees Celsius) at admission had 78% decreased likelihood of death compared to patients with abnormal body temperature (aOR, 0.22; 95% CI, 0.05-0.99; p-value, 0.049), hyperthermia is a sign of severe COVID 19 and depending on the grade of severity, the more severe the disease is the higher the mortality. Physiologically, fever is caused by cytokines that alter the hypothalamic temperature set point in febrile illnesses. Various cytokines and interleukins are produced in COVID 19 disease with a peak at approximately 10 days of symptom onset causing a cytokine storm that leads to the most severe form of COVID 19 associated with hyperpyrexia, systemic inflammatory response syndrome, multiple organ dysfunction and very high mortality.

About half (48%) of the patients had low oxygen saturation (SPO2 <92). Normal oxygen saturation (SPO2 of 92-100%) throughout the period of admission was associated with 89% decrease in the likelihood of death (aOR-0.11; 95% CI 0.03-0.44; p-value-0.002), on multivariate analysis, this finding was consistent with that in Mulago [15]. COVID 19 causes pneumonia and impairs gaseous exchange in the lungs necessitating oxygen supplementation and without oxygen patients deteriorate quickly and die. In Uganda this was mostly during the second wave where delta variant caused very severe disease that overwhelmed the health system noticeable by many deaths, breakdown in the oxygen plants in many CTUs across the country due to overuse. When the oxygen plant for LRRH broke down, at the same time with that of Soroti and Gulu regional referral hospitals, Oxygen could only be got from Jinja with a lot of delay. This even worsened the situation.

34% had tachypnea RR > 18 breaths per minute and 66% had normal respiratory rate, RR< 18breath per minute on admission, normal respiratory rate was found to be associated with decrease likelihood of death at bivariate analysis but there was no association at multivariate analysis, this could be because of the limited sample size or effect of co-founding because patients with normal oxygen saturation have normal respiratory rate and normal oxygen saturation was found to be associated with reduced likelihood of death. Physiologically the respiratory rate increases when the blood oxygen concentration falls below normal (SPO2<92%) to compensate for the low SPO2, however this compensatory mechanism fails in severe hypoxia. 37% patients had tachycardia (PR > 100bpm), a non specific sign and it was found not to be associated with mortality, it can be caused by anything including anxiety which is common in COVID 19 patients immediately after learning they are suffering from the disease. About 20% of the patients had consistently measured hypertension (BP > (140/90) mmHg).

### The role of co-morbidity

The most common co-morbidity among patients was hypertension (30%) of cases which was associated with increased mortality at bivariate but not at multivariate analysis as opposed to other studies that indicated hypertension increases mortality in COVID 19 [27] [21] [28]. Diabetes mellitus was in (19%) of cases, it was associated with increased mortality at bivariate but not multivariate analysis as opposed to other studies [27] [21] [28]. COVID19 has been found to worsen DM and make control of hyperglycemia difficult in DM; many of the DM patients with COVID 19 come in emergency with acute complications of DM like DKA, HHS. COVID 19, studies have showed that COVID 19 directly damages beta cells of the islet of langerhans of the pancreas directly causing DM. DM has been found to worsen COVID 19 [29] [30]. HIV was in 10% of the total number of patients admitted with COVID 19 but it was not associated with mortality both at bivariate and multivariate analysis as opposed to other studies that showed that HIV is associated with increased mortality [31] [32] [33]. Current or concomitant Malaria infection with COVID 19 co-infection occurred in 9% of admitted patients, Malaria was not associated with mortality unlike study of Malaria consortium in multiple COVID 19 treatment centers in Uganda that reported Malaria is protective of COVID 19 [34]. The rest of the co-morbidities were few and not associated with mortality in this study, most of these co-morbidities were not associated with mortality at multivariate level probably because of the limited sample size that could have lowered their power of association, especially for hypertension, DM and HIV, or because of the effect of co-founding because most of these patients with co-morbidity and advanced age suffered severe COVID 19 with low oxygen saturation < 92% either in room air or on oxygen supplementation which is associated with increased mortality in other studies.

Chronic liver disease was associated with mortality even when it was not among the common comorbidity, patients without chronic liver disease had 99% decreased mortality (aOR-0.01, 95% CI 0.001-0.46, p-value-0.017), this is in line with other studies [35] [36]. COVID 19 worsen liver disease by a number of mechanisms, among them are direct damage of the hepatocytes by SARS Cov 2 virus, host inflammatory response to SARS Cov 2, drug induced liver injury during treatment of COVID 19 that all worsen liver function. Chronic liver disease also worsens COVID 19 by lowering the immunity.

## Strength of the study

The study period was long enough from May 2020 to March 2022, this was about the time the first and last patients were admitted to LRRH-CTU taking into account all the three waves of COVID 19 therefore eliminating selection bias and hence better generalization of the findings.

This study also excluded asymptomatic patients who were admitted to prevent community spread of COVID 19; those who were studied were actually sick patients that needed treatment and in-hospital care. Therefore the outcome gives a better picture of the capacity of the hospital in managing COVID 19.

There was no bias since all the patients who were admitted in the CTU were considered for selection, only those who did not meet the eligibility criteria were left out.

## Limitations of the study

Being a retrospective study our study;

Because we collected data retrospectively, data in the chart may have not been accurate all the time. To go around this, the researcher who is highly skilled had to look through this charts himself in order to understand the medical terms and processes and try to make sense of them as accurately as possible.

Some charts did not have all the required information and were not considered in the study, to go around this problem of missing data; a large sample size was used.

The sample came from a very highly selected group of patients since most of the patients who were admitted in the second and third wave were very sick (moderate to severe) meeting defined criteria for admission, and Due to all these factors, generalization of findings was difficult.

## Conclusion

The in-hospital mortality rate in our cohort of COVID 19 patients admitted at LRRH was high. Not having chronic liver disease, normal oxygen saturation, normal body temperature and younger age were associated with decreased likelihood of death.

## Supporting information

COI disclosure forms

## Data Availability

All data produced in the present study are available upon reasonable request to the authors

## Ethical Approval

The Gulu University Research Ethics Committee (GUREC) approved this protocol and exempted written consent. All patient data were handled anonymously and strictly confidential.

## Transparency Declaration

The lead author affirms that this manuscript is an honest, accurate, and transparent account of the study being reported; no important aspects of the study have been omitted; any discrepancies from the study as planned (and, if relevant, registered) have been explained.

## Patient and public involvement

Patients or the public were not involved in the design, conduct and dissemination of this research.

## Data sharing

All the data relevant to the study are all included in the article or uploaded as supplemental information and this is an open access article provided the original work is not commercially used.

## Acknowledgements

Our appreciation to the staffs of Lira regional referral hospital for allowing us to conduct this research, the students and staffs of Lira University for their support and guidance and the COVID 19 health care workers that managed COVID 19 patients.

---

**Figure 1.**
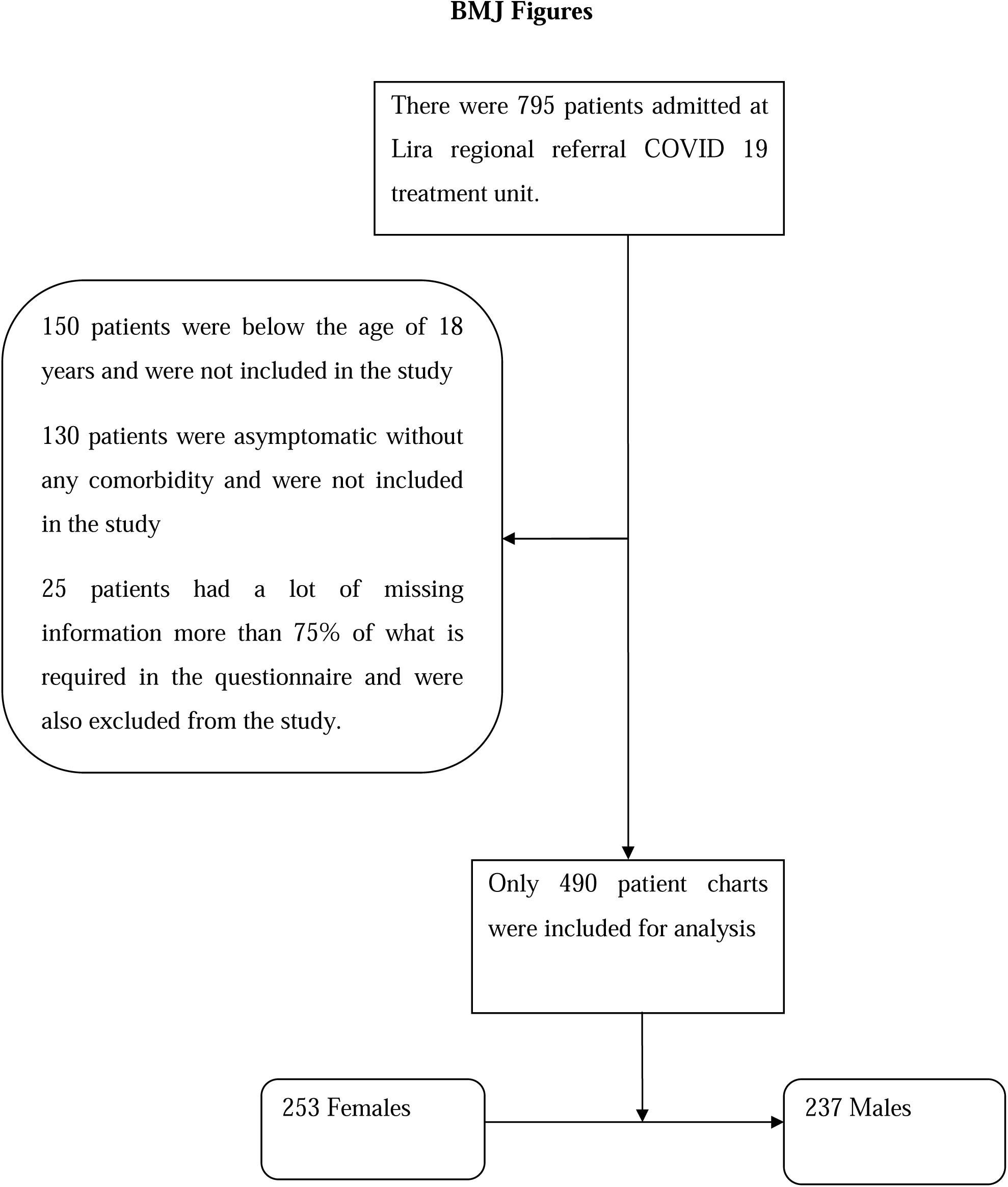

